# A highly divergent SARS-CoV-2 lineage B.1.1 sample in a patient with long-term COVID-19

**DOI:** 10.1101/2023.09.14.23295379

**Authors:** Elena Nabieva, Andrey B. Komissarov, Galya V. Klink, Stanislav V. Zaitsev, Maria Sergeeva, Artem V. Fadeev, Kseniya Komissarova, Anna Ivanova, Maria Pisareva, Kira Kudrya, Daria Danilenko, Dmitry Lioznov, Ryan Hisner, Federico Gueli, Thomas P. Peacock, Cornelius Roemer, Georgii A. Bazykin

## Abstract

We report the genomic analysis of a highly divergent SARS-CoV-2 sample obtained in October 2022 from an HIV+ patient with presumably long-term COVID-19 infection. Phylogenetic analysis indicates that the sample is characterized by a gain of 89 mutations since divergence from its nearest sequenced neighbor, which had been collected in September 2020 and belongs to the B.1.1 lineage, largely extinct in 2022. 33 of these mutations were coding and occurred in the Spike protein. Of these, 17 are lineage-defining in some of the variants of concern (VOCs) or are in sites where another mutation is lineage-defining in a variant of concern, and/or shown to be involved in antibody evasion, and/or detected in other cases of persistent COVID-19; these include some “usual suspects,” such as Spike:L452R, E484Q, K417T, Y453F, and N460K. Molecular clock analysis indicates that mutations in this lineage accumulated at an increased rate compared to the ancestral B.1.1 strain. This increase is driven by the accumulation of nonsynonymous mutations, for an average dN/dS value of 2.2, indicating strong positive selection during within-patient evolution. Additionally, there is reason to believe that the virus had persisted for at least some time in the gastrointestinal tract, as evidenced by the presence of mutations that are rare in the general population samples but common in samples from wastewater. Our analysis adds to the growing body of research on evolution of SARS-CoV-2 in chronically infected patients and its relationship to the emergence of variants of concern.

## Introduction

Although COVID-19 is typically an acute disease lasting days or weeks, rare cases of persistent SARS-CoV-2 infection are known, and these are usually associated with suppressed host immune systems^1–5^. Mutation accumulation continues throughout the course of the disease, and is in fact faster than in inter-person transmission ^5–7^. It is hypothesized that this long-term mutational accumulation under weakened immune pressure can sometimes allow the virus to evolve the sorts of beneficial combinations of mutations that give rise to novel strains with immune evasion potential, and that this mechanism could explain the emergence of VOCs on long phylogenetic branches^8–10^.

## Results and Discussion

### Case description

The female patient was admitted to the hospital in the city of Kaluga, Russia in September 2022 with symptoms of irritable bowel syndrome, including diarrhea and borborygmus. She had not been vaccinated against COVID-19. Initial screening tests revealed positive results for both SARS-CoV-2, detected by RT-PCR from a nasopharyngeal swab, and HIV, detected by an antigen/antibody HIV assay. The patient (henceforth referred to as “patient K” for “Kaluga”; the identifier is not known to anyone outside the research group and is only used for purposes of this publication) claimed that she had been HIV-positive since 2005 and never received HAART. The patient also had a dry cough and fatigue but was afebrile. Over the course of her hospital stay, the patient was tested for SARS-CoV-2 by RT-PCR four more times (on days 3, 6, 9, and 13 post-admission), with all results being positive. She has had no recent travel history.

Despite the COVID-19 diagnosis, the patient exhibited no signs of lung injury on X-ray imaging and did not require oxygen support. However, her complete blood count was remarkable for leukopenia (1.6-2.2%) and a low hemoglobin level (86-108 g/l). On day 14 post-admission, the patient refused any further medical treatment and discharged herself from the hospital.

In April, 2023 the patient was admitted to a hospital with diarrhea, myalgia, borborygmus and progressing fatigue.

### A SARS-CoV-2 sample on an ultra-long branch

We sequenced the COVID-19 sample from patient K and deposited it in GISAID as hCoV-19/Russia/KLU-RII-MH107906S/2022. Bioinformatic analysis of the sequence strongly suggests that it comes from a case of ultra-long infection. Specifically, the sample is identified as belonging to the B.1.1 lineage^11^. The B.1.1 lineage has become nearly extinct by late 2022: besides the variant reported here, it comprised just 0.004% (69 out of 1,608,995) of GISAID sequences with “Collection date” later than September 1, 2022 (GISAID queried on 2023-03-11), and was absent (0%) among the 16,462 sequences collected at that time in Russia. To verify the phylogenetic placement of the sample in the B.1.1 clade, we considered alternative tools: pangoLEARN^12^ and NextClade^13^. The latter uses an approach similar to that of UShER, while the former employs an entirely different machine-learning method. Both agreed that the most likely placement is B.1.1.

Despite the robust placement of the sample in the early B.1.1 clade, its lineage is highly divergent, with 89 point mutations. Meanwhile, the closest neighbor in the phylogenetic tree built by UShER using the 14,255,578 GISAID, GenBank, COG-UK, and CNCB sequences (accessed on 2023-03-12) is hCoV-19/Russia/MOW-CSP-KOM-0303_S25/2020, collected in Moscow, Russia on 2020-09-23 (Fig. 1). Out of the 10 phylogenetically adjacent samples, 9 were obtained in Russia between 2020-07-16 and 2021-01-07 (Fig. 1), confirming that the lineage likely resulted from a community infection around the estimated time of the internal node. According to Bayesian estimation with BEAST2^14^, the median age of the patient K branch was 2.06 years, putting the date of its ancestor on 2020-09-10 (consistent with the dating of the nearest outgroup sequences), although the 90% HPD was very high (0.0001 to 23.4 years). Together, these observations strongly suggest that this is indeed a case of persistent (up to two years or more) SARS-CoV-2.

**Figure 1.**
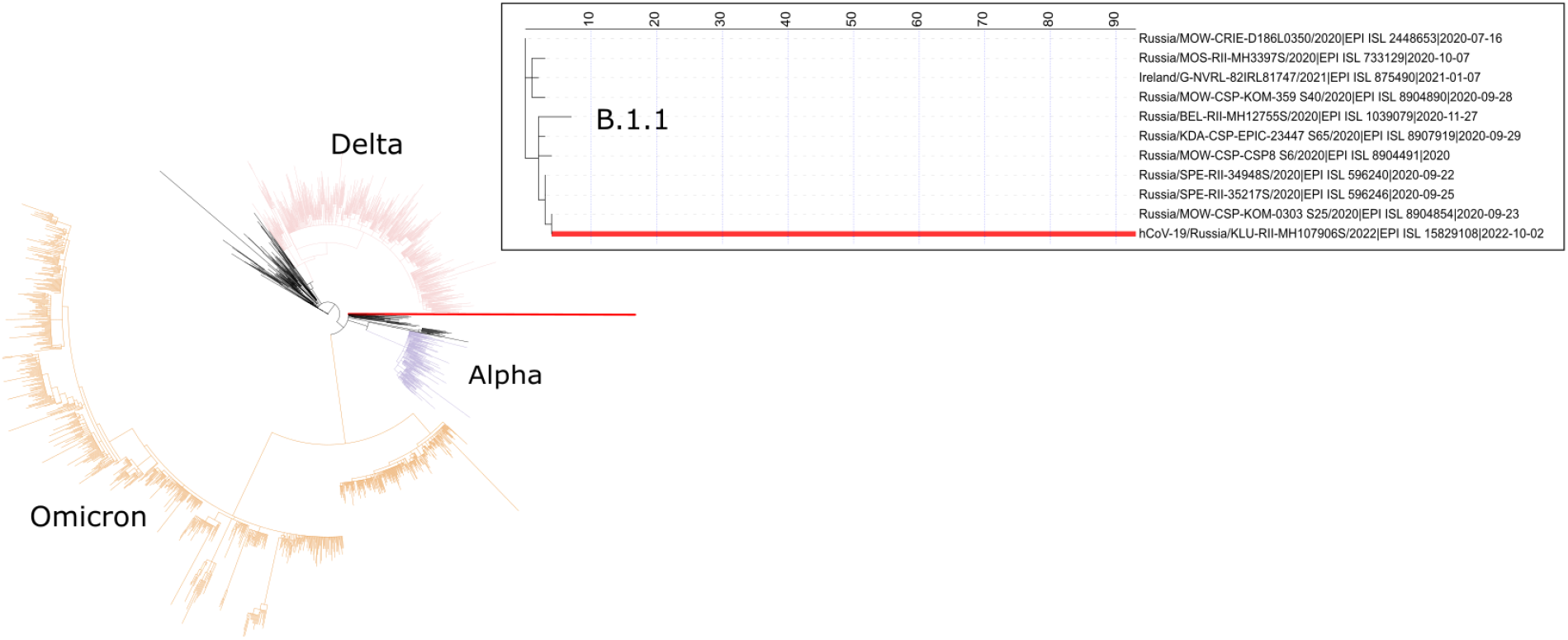
Placement of hCoV-19/Russia/KLU-RII-MH107906S/2022 in the phylogenetic tree by UShER^11^, with population downsampled to 2000 sequences. The inset shows the local phylogenetic vicinity of the patient K sample (tree visualization by IToL^15^).

### Long-term infection most likely explanation for the number of mutations

In the absence of time-course samples from the patient, we establish long-term infection as the most likely explanation for the high number of mutations observed by ruling out the two alternative possibilities that could also explain the “long branch” observation, namely, contamination and recombination.

We rule out contamination by observing that the sample does not have an excess of variants at intermediate read frequencies. In particular, of 104 nucleotide point mutations and indels having read fraction > 0.5 (our definition of consensus mutations), just a few are at intermediate read frequencies: only 10 have read fraction < 0.8, and of these, only 2 have read fraction < 0.6. By the same reasoning, we do not observe a significant intrahost variation in the sample, although this of course is based on just a single sample and does not exclude the possibility of additional variation within the host which is frequently observed in persistent infections ^5,16^.

To see if the observed sequence may be a result of recombination event(s), we ran it through the tool sc2rf (https://github.com/lenaschimmel/sc2rf) to look for evidence of recombination between CoVariants clades (https://covariants.org). No such evidence was observed. As an alternative approach, we used RIPPLES^17^ which also detected no recombination events. The “interleaving” of mutations from different VOCs and non-VOC mutations, as well as the presence of non-VOC derived alleles in sites of VOC-defining mutations (Table 1), also contradict the hypothesis of the sequence resulting from recombination event(s) involving phylogenetically distant lineages.

**Table 1.** Select Spike mutations in the patient K sample.

| Mutation | VOCs with the same mutation or another mutation at the same site in their origin | Immune evasion | Chronic infection | In wastewater | Other notable lineages with the same mutation or another mutation at the same site at their origin |
| --- | --- | --- | --- | --- | --- |
| T19A | *Delta\**, BA.2 |  |  |  |  |
| S50L |  |  | 5 |  |  |
| T95I | BA.1 |  | 2, 1 |  |  |
| R346K |  | 28, 29, 30 |  |  | BA.1.1, *XBB*, *BQ.1.1* |
| R403K |  | 31 |  |  |  |
| K417T | Gamma, *Beta*, *BA.1*, *BA.2* | 32, 33, 31, 34 |  | 24 | BA.2.38 |
| K444T |  | 30 |  | 24 | BQ.1 |
| Y449H |  | 35 |  | 24 |  |
| L452R | Delta | 32, 31, 36 | 1, 5 |  | BA.4, BA.5, BQ.1, *BA2.12.1* |
| Y453F |  | 34 | 5 |  | mink cluster V |
| N460K |  | 32, 30 |  | 24 | BA.2.75, BQ.1, XBB |
| A475S |  | 31 |  |  |  |
| S477N | BA.1, BA.2 | 37 |  |  |  |
| E484Q | *Beta*, *Gamma*, *BA.1*, *BA.2* | 32, 33, 31, 35, 34 | 2, 1 | 24 | Kappa |
| Q498H | *BA.1*, *BA.2* |  | 1 | 24 |  |
| P621S |  |  | 5 |  | XAY |
| N764K | BA.1, BA.2 |  |  |  |  |
| Δ141-144 | Alpha (Δ144), BA.1 (Δ142-144) | 25 | 2, 6, 5 |  |  |
| Δ210 |  | 25 |  |  |  |

The table lists the mutations that are lineage-defining in some of the variants of concern (VOCs) or are in sites where another mutation is lineage-defining in a variant of concern, and/or shown to be involved in antibody evasion, and/or detected in other cases of persistent COVID-19.
\*Lineages where different mutations at the same site were found in the ancestor, or papers mentioning different mutations in the same site are italicized.
Other mutations in the S gene are: H49R, H66Q, G72R, V126A, G181E, F186I, L216F, A222D, Y248H, Q271R, A372T, R765L, V772A, T1006I, A1078S, D1165E

### Elevated mutation accumulation rate in the patient K sample

We plotted the number of mutations accumulated since divergence from the Wuhan ancestral sequence for the patient K sample and sequences from the UShER-selected subtree containing it ^11^ (Fig. 2) using TempEst ^18^. One can see that the rate of mutation accumulation “jumps” dramatically in the beginning of 2022, corresponding to the disproportionately large number of changes at the origin of the Omicron variants ^19,20^. Remarkably, the patient K sample (red dot) falls on the higher end of the Omicron cloud corresponding to the “BA.2” samples obtained around the same time. Thus, the rate of mutation accumulation in the lineage of the patient K sample has been comparable to the rate at the origin of the Omicron clade. Notably, while the rates are similar, the identity of the accumulated mutations is different: the patient K sample shares just a few mutations with BA.2 (see below).

**Figure 2.**
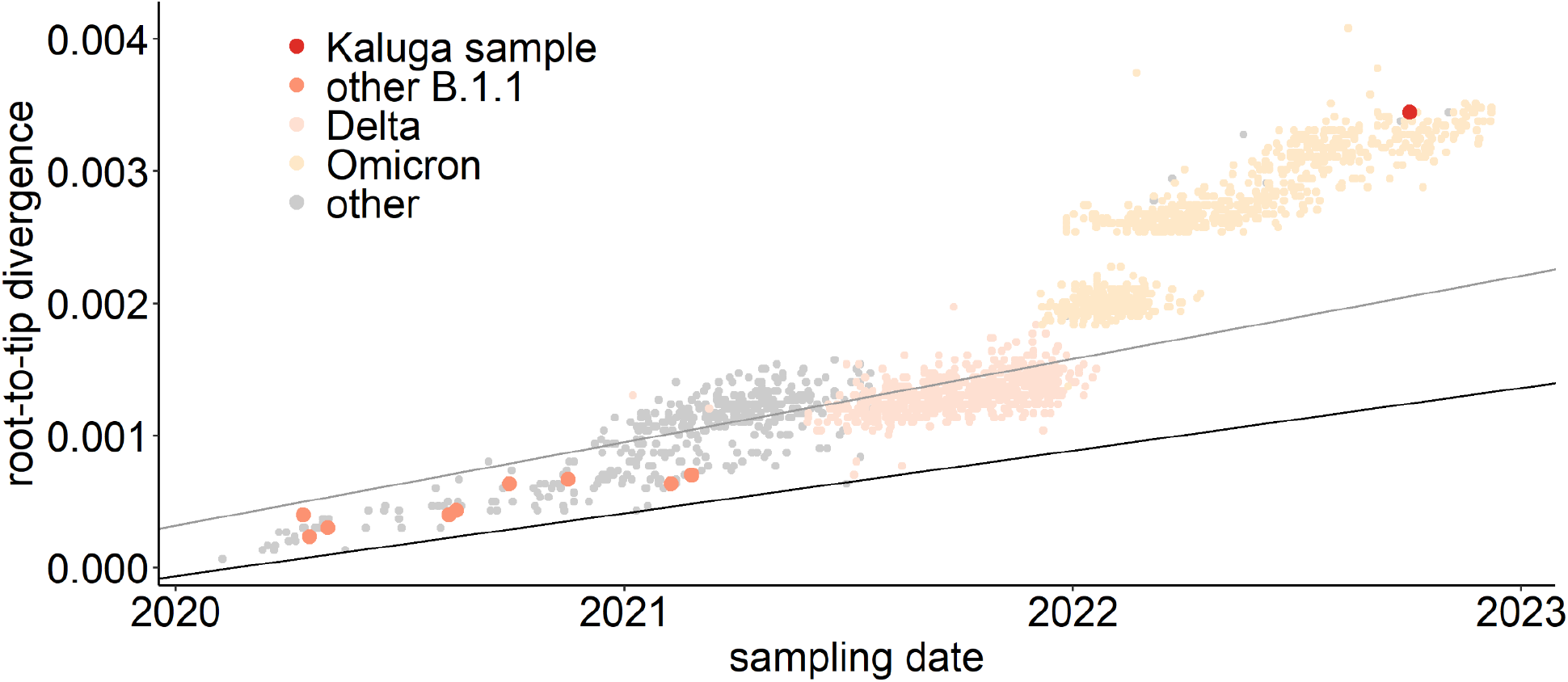
Regression of root-to-tip distance on sampling date. The regression lines are calculated for all non-Omicron, non-patient K samples (slope = 6.3x10^−4^) and just for the B.1.1 samples (slope = 4.7x10^−4^). The patient K sample is high above both regression lines, indicating accelerated evolution at this lineage.

83% of the single-nucleotide substitutions were non-synonymous, similar to the values observed for samples from immunocompromised patients and from ancestral branches of VOCs^2^. To evaluate whether the rapid evolution of the patient K sample was caused by positive selection, we applied BUSTED ^2^ method implemented in HyPhy^22^. BUSTED tests whether a sequence experienced positive selection on specified branch(es) of a phylogenetic tree. We performed separate analyses for the entire concatenated coding sequence (excluding nucleotide positions with overlapping reading frames), for Spike gene alone, and for the entire coding sequence with Spike gene removed. For the entire coding sequence and for spike, we detected positive selection on the branch leading to the patient K sample (p = 0.04 for concatenated genes and p = 0.0034 for Spike gene). For concatenated genes, the relative rate of non-synonymous evolution (dN/dS ratio) was 2.2 for the branch leading to the patient K sample and 0.6 for the rest of the tree of B.1.1 samples. For the Spike gene, these values were 5.7 and 0.7, respectively. The positive selection (dN/dS > 1) observed for the patient K branch contrasts with the predominantly negative selection (dN/dS < 0.5) observed in the global evolution of SARS-CoV2^23^. For concatenated genes without Spike, there was no evidence of positive selection on the patient K branch (p = 0.2370). Therefore, an increase in mutation accumulation rate on the patient K branch was likely driven by positive selection specific to this branch, acting (mainly) on the mutations in the Spike protein.

### Mutations in the Spike protein are typical for long-term infection, immune escape, and VOCs

Of the 89 private point mutations in the sample, 33 occur in the Spike gene, with an additional two inframe deletions (Table 1, Fig. 3). These include 17 point mutations that have been seen in Variants of Concern or in sites where other mutations have been found in Variants of Concern, and/or in immunocompromised patients, and/or are associated with immune escape. Among these mutations are L452R, E484Q, K417T, N460K, and Q498H. Not just the occurrence, but also the combinations of some of these substitutions parallel lineage-defining mutations in Variants of Concern: T95I, S477N, N764K co-occur in BA.1 (with K417T, E484Q, Q498H occurring in sites of other lineage-defining BA.1 mutations), while K444T and L452R, and N460K co-occur in BQ.1. Additionally, the sequence also has several ORF1a mutations that are associated with hypermutated, chronic-infection sequences, most notably T1638I (NSP3:T820I), K1795Q (NSP3:K977Q)^1^; the latter mutation has additionally been identified in a number of NYC wastewater cryptic lineages, in several of which its fraction was close to 100% ^24^. These mutations may constitute the response of the virus to selective pressure from the host immune system.

**Figure 3.**
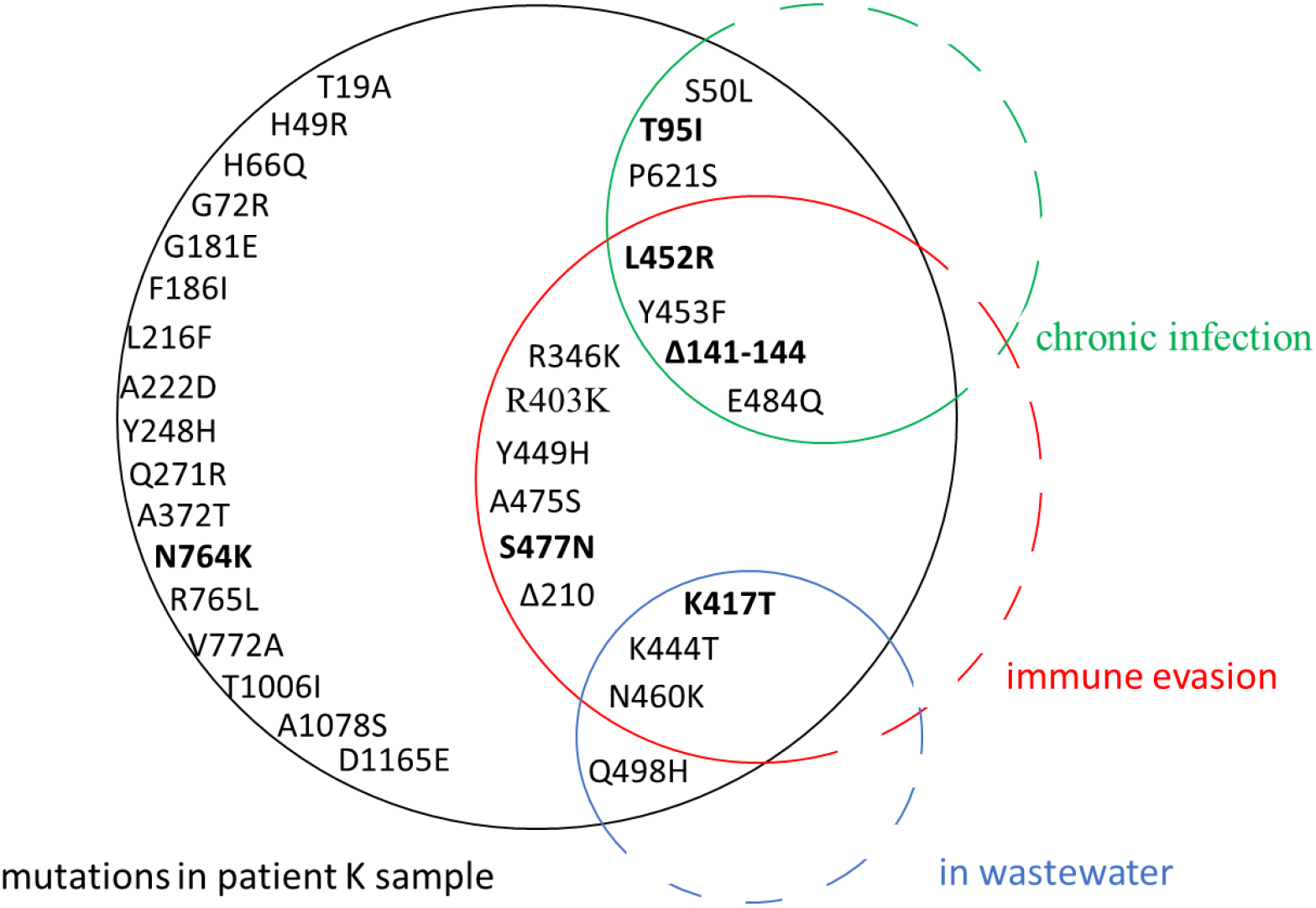
A schematic representation of the spike mutations in the patient K sample and their overlap with previously described mutations found in chronic infections, associated with immune evasion, or overrepresented in wastewater samples (see Table 1 for corresponding references). Mutations that were also observed in the ancestors of VOCs are in bold.

In addition to the point mutations, the sample also contains two deletions in the S gene: Δ141-144 and Δ210. They occur in two of the four “Recurrent Deletion Regions” in the NTD (RDR2 and RDR3) in the McCarthy et al.^25^ nomenclature and map to distinct antibody epitopes^1^ and overlap or are adjacent to deletions in VOCs: Alpha (Δ144) and Omicron/BA.1 (Δ211) ^26^.

Interestingly, the Y453F+Δ141-144 combination has been seen also in early (but not late) samples from a case of persistent COVID-19 in a lymphoma patient lacking humoral immunity due to treatment with rituximab ^5^. Similar deletions have been shown to arise recurrently in chronic infections ^1^.

In addition to these acquired mutations, there is a T14408C nucleotide substitution that reverses the Orf1b:P314L (RdRp:P323L) mutation present in 98% of sequences that carry Spike:D614G. RdRp:P323L may stabilize the RdRp^27^. Based on the GISAID data, this reversion is shared with the phylogenetic neighbor Russia/MOW-CSP-KOM-0303_S25/2020; if the reversion in this neighbor is genuine, this means that the reversion has in fact happened prior to the long-term infection.

### To the gastrointestinal tract and back again?

The pattern of Spike receptor binding domain (RBD) mutations in this sequence shows remarkable overlap with the set of mutations that have been repeatedly found in mutations from “cryptic lineages” detected in wastewater^24^, which are hypothesized to be long-term chronic infections with a gastrointestinal tropism. Some of them are also found in VOCs, e.g., K417T (Gamma), K444T (BQ.1), and N460K (BA.2.75, BQ.1, XBB), while others have been found recurrently in wastewater but otherwise are rare among sequences in GISAID. One example is Spike:Q498H, which is found in only 96 sequences in GISAID (accessed on December 22, 2022), but found in 7/9 sewersheds studied in ^24^ (a different substitution at the same site, Q498R, is present in a number of Omicron lineages and therefore widespread). This is consistent with the fact that the patient has presented with gastrointestinal (GI) symptoms.

Moreover, of particular interest is the double mutation in codon 828 of Spike (CTT-><u>T</u>T<u>G</u>). Conceivably, during the presumed long intrahost evolution, the sequence could go through the intermediate state of TTT (Spike:L828F) which was later followed by the F828L reversion to a different leucine codon. L828F is remarkable in that it comes up repeatedly in cryptic lineages found in wastewater, while being extremely rare in general SARS-CoV-2 sequences (almost exclusively sampled from the respiratory tract)^24^. We propose that L828F may represent a specific adaptation to the GI tract during chronic infection. Based on this proposed chain of events and on the overlap with other mutations recurrently found in wastewater samples described above, we suggest that the virus has resided within and adapted to the patient’s GI tract during some period of time in the long course of the infection. Eventually, though, the 828L reversion brought it back to the consensus sequence presumably more adapted to nasopharyngeal localization, and the virus was ultimately detected in the nasopharynx. Admittedly, this suggested mechanism is highly speculative: the 828F variant by itself was not observed in the patient K, the double CTT-><u>T</u>T<u>G</u> mutation could have proceeded through the synonymous (L) CT<u>G</u> rather that the nonsynonymous (F) <u>T</u>TT intermediate, and the pathway for reverse transfer of the GI-adapted variant into nasopharyngeal localization is unclear. Still, this finding adds further, albeit circumstantial, evidence to the idea that a subset of chronic infections may be GI (likely accounting for cryptic wastewater lineages). At present, it is thought that such GI chronic infections likely represent an evolutionary dead end due to scant evidence of transmission of these lineages. However, in this sequence, we find tentative evidence that this virus has re-adapted back to the respiratory tract, giving a potential mechanism for how such a virus could re-evolve to become transmissible by the airborne route.

### Impaired growth in cell culture compared to the B.1 ancestor

In an attempt to find particular markers of phenotypic adaptation, we isolated the virus (KLU) from the patient K swab specimen and compared the growth rate of the KLU virus and ancestral B.1 strain in the Caco-2 human intestinal epithelial cell line (ATCC, Cat # HTB-37). At first we checked three different doses (0.1, 0.01 and 0.001 TCID50/cell) and two incubation temperatures (34°C and 37°C). The temperature dependence of the growth rate was the same for both viruses indicating no loss of adaptation to the upper respiratory site of infection for the KLU virus (Fig. 4A). Secondly, we checked three different pH of the culture media: neutral pH = 7.5, acidic pH = 6.5 and alkaline pH = 8.5. The pH dependence of the growth rate was the same for both viruses with no difference between neutral and low pH and impaired growth at higher pH, indicating no adaptation of the KLU virus to the potentially different environment of the gastroenterological tract (Fig. 4B). For both viruses, no cytopathic effect was observed at any time point. At the same time, we found that the KLU virus growth was overall significantly impaired in comparison to the B.1 strain in the Caco-2 model cell line. The observed difference was similar to that described earlier for the Omicron strains and can be linked to less effective antagonizing of the interferon response in human cells due to mutations in particular viral proteins^38^.

**Figure 4.**
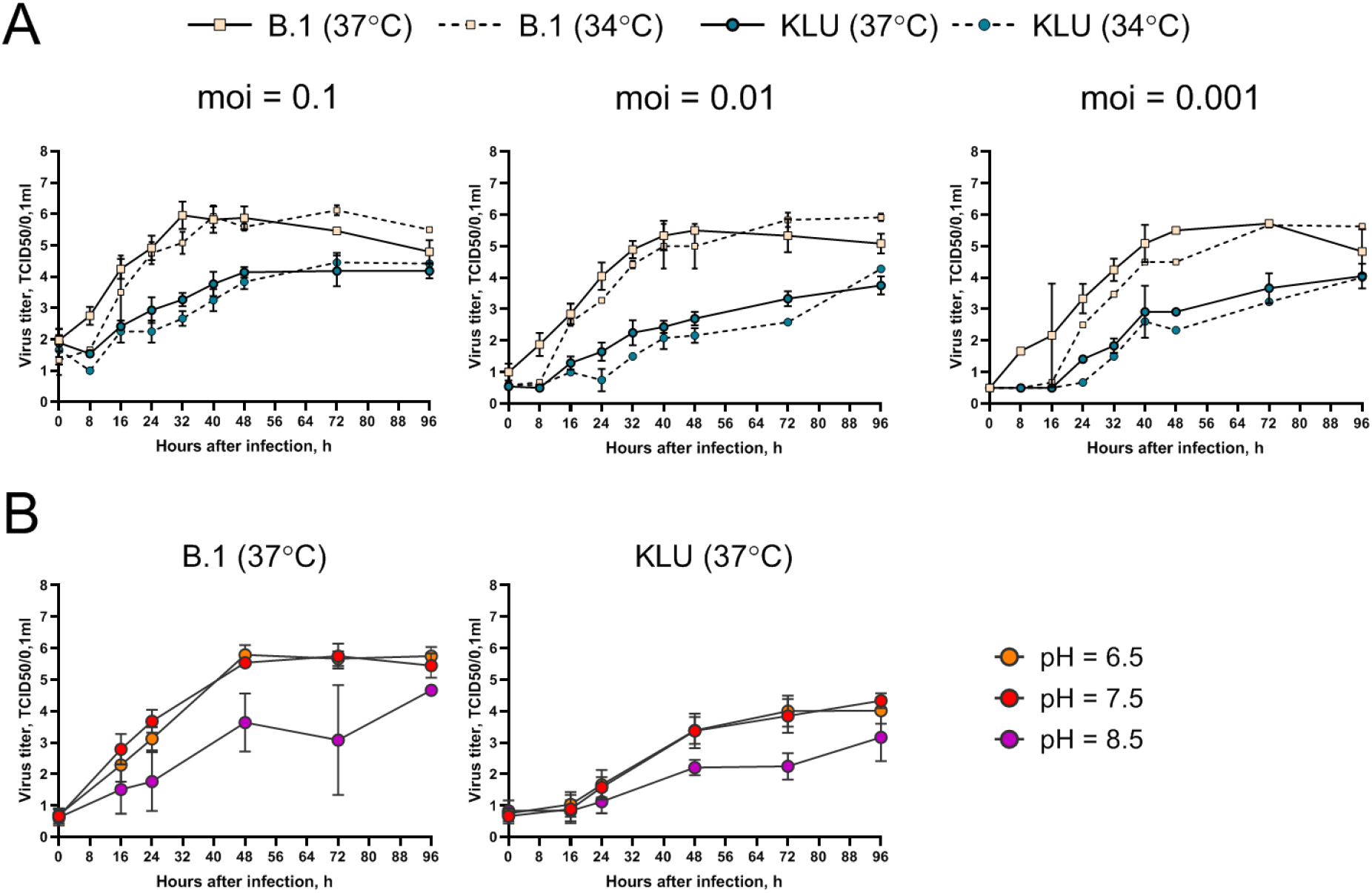
Growth curve of the KLU virus and the B.1 ancestral strain in Caco-2. (A) Cells were infected at indicated moi (TCID50/cell) and incubated at 34 or 37°C. (B) Cells were infected at moi = 0.01 TCID50/cell and incubated at 37°C in the culture medium with indicated pH. Samples of culture medium taken with an 8-24 hour interval after inoculation were titrated for virus infectious activity in the VeroE6/TMPRSS2 cell line.

### Is there risk of a new VOC?

There is growing support for the hypothesis that at least some VOCs, such as Alpha and Omicron, have evolved in patients with chronic infection and escaped into the general population ^8–10^. It has been shown that many of the lineage-defining mutations in VOCs are associated with antibody escape (providing selective advantage in the largely immune-competent population in the later stages of the pandemic), but that at least some of them reduce RBD affinity for ACE2, an effect that is rescued in BA.1 by the compensatory mutations Q498R and N501Y^39^. Neither is observed in our sample, although, as we discuss above, it contains a different mutation in the 498 codon, namely, Q498H. The rarity of Q498H in GISAID suggests that it may not be responsible for airborne spread, although it has been experimentally shown to increase the ACE2 affinity on non-N501Y spike background^40^. Furthermore, the Y453F mutation greatly increases the ACE2 affinity in both BA.1 and BA.2, as well as Wuhan Hu-1 backgrounds, while R403K substantially increases ACE2 affinity on a BA.2 spike background, but not a BA.1 or Wuhan Hu-1 spike background^40^. Since this sample shares mutations with both BA.1 and BA.2, there is a theoretical possibility of its increased infectivity.

As of March 14, 2023, the hCoV-19/Russia/KLU-RII-MH107906S/2022 sample remains the sole representative of this lineage in GISAID, although GISAID carries 9,003 additional samples from Russia, including 195 from Kaluga region, with later sampling dates. Together with the fact that the virus has been sampled from the likely source of long-term evolution (the HIV+ patient), this suggests that this lineage has not led to substantial onward transmission. Still, it indicates that SARS-CoV-2 has not yet run out of opportunities for further adaptation.

## Conclusion

We have described a case of a likely long-term infection in an HIV+ patient. Sampled in 2022, it is most confidently placed in the B.1.1 clade that was largely extinct by the time. As just a single sample from this patient is available, the persistence of the virus cannot be verified by repeated positive PCR tests throughout the presumed duration of infection. Instead, the evidence for persistent infection is circumstantial, and is based on the likely immunosuppressed (HIV+) status of the patient together with the phylogenetic placement of this sample. In addition to the long branch from the putative root indicative of a long-term infection, remarkable features of the sample include the presence of mutations defining several VOC lineages and/or associated with immune evasion, and an apparently elevated mutation accumulation rate. In fact, the rate is similar to that at the origin of the Omicron clade, suggesting that the evolutionary processes that shaped its evolution may be similar to those that shaped the evolution of the Omicron variants and, conversely, lending further support to the hypothesis that VOCs such as Omicron emerged in the course of chronic infection^39^. The increased fraction of nonsynonymous mutations indicates strong positive selection in this lineage, particularly in the spike protein.

An intriguing feature of the patient K sample is the presence of several mutations that have been found almost exclusively in wastewater lineages. This observation suggests the possibility of persistence for some period in the GI tract. The connection between immune-evading long-term infection, gastrointestinal symptoms, and the risk of emergence of VOCs suggested by the observations in this sample highlight the need for wastewater monitoring.

## Methods

### Ethics declaration

The sample used in this study was collected as part of the ongoing surveillance of SARS-CoV-2 variability routinely conducted at the laboratories of the CoRGI consortium (CoRGI). Genomic epidemiology study conducted by CoRGI consortium was reviewed and deemed exempt by the Local Ethics Committee of Smorodintsev Research Institute of Influenza (protocol No. 152, 18 June 2020). This case report involved a retrospective retrieval, review and analysis of clinical data. These activities were approved by the Local Ethics Committee of Smorodintsev Research Institute of Influenza (protocol No. 203, 18 May 2023) in accordance with the ethical standards laid down in the 1964 Declaration of Helsinki and its later amendments. The patient gave written informed consent prior to sample collection, according to CARE guidelines (See: https://www.care-statement.org/). To maintain patient privacy, all identifying information has been removed from the publication.

### Sample collection

Nasopharyngeal swab was collected in virus transport medium. Total RNA was extracted using Auto-Pure 96 Nucleic Acid Purification System (Allsheng, China) and NAmagp DNA/ RNA extraction kit (Biolabmix, Russia). Extracted RNA was immediately tested for SARS-CoV-2 using Novel Coronavirus (2019-nCoV) Nucleic Acid Diagnostic Kit (Sansure Biotech, China).

### Sequencing

Whole-genome amplification (WGA) of SARS-CoV-2 virus genome was performed using Midnight 1200bp amplicon primer set^41^ and BioMaster RT-PCR-Premium Kit (Biolabmix, Russia). Library preparation was performed with Fast PCR-FREE FS DNA Library Prep Set (MGI, China). Finally, sequencing was performed on MGI DNBSEQ-G400 using FCL SE100 high-throughput sequencing set.

### Bioinformatic analysis

Positive selection was tested with the BUSTED^21^ method of the HyPhy package^22^. To obtain the test dataset, we merged B.1.1 samples that differed in less than 5 positions with cd-hit^42^, took a random subset of 500 samples, removed samples with premature stop codons, added the patient K sample and reconstructed a phylogenetic tree with IQTree ^43^.

The age of the patient K lineage was estimated based on the analysis of twelve B.1.1 sequences of the Spike gene obtained as a clade of the tree constructed for testing for positive selection, consisting of the patient K sample and samples that were closest to it. The age was estimated with BEAST2 v.2.7.4 ^14^ with the following parameters: HKY substitution model; gamma site model with 4 rate categories; coalescent tree prior with constant population; random local Log Normal clock model (M=7.0E-4, S=1.25, mean in real space); 500 mln MCMC chain length; other parameters were default for BEASTv.2.7.4.

### Virus isolation and growth curve

Live virus (KLU) was isolated from the patient K swab samples in VeroE6/TMPRSS2 cells (JCRB, #JCRB1819). Culture was inoculated for 2 hours with swab material diluted 1/10 in DMEM (Biolot) supplemented with 2% heat-inactivated fetal bovine serum (SC, Biolot), 1% anti-anti (Gibco) and then incubated for 4 days until 70% cytopathic effect. The sample was subsequently passaged one time in VeroE6/TMPRSS2 cells to generate the working stock.

Growth curve experiments were performed in Caco-2 cells (ATCC, #HTB-37). KLU virus was compared with the B.1 strain hCoV-19/St_Petersburg-3524V/2020 virus (GISAID EPI_ISL_415710). Cells were grown until 100% monolayer in 12-well plates in MEM culture medium (Gibco) supplemented with 20% FBS (Gibco), 1% GlutaMax (Gibco), 1% sodium pyruvate (Gibco) and 1% NEAA (Biolot). Monolayer cells were inoculated with a particular viral dose (0.1, 0.01 or 0.001 TCI50/cell) in 0.5 ml neutral medium with 5% FBS for 1 hour at 37°C, then inoculum was removed and 3 ml medium with 5% FBS was added. The neutral (original) medium pH was 7.5 at 37°C. To get the low pH medium we added 1M MES (Amresco) until pH = 6.5; to get the high pH medium we added 1M Tris-base (Amresco) until pH = 8.5. Samples of culture supernatant were taken at 8-24 hour intervals after inoculation and titrated for virus infectious activity in the VeroE6/TMPRSS2 cell line by standard TCID50 method.

## Data Availability

All data produced in the present work are available in GISAID with the identifier EPI_ISL_15829108.

https://gisaid.org/

## Data Availability

The consensus sequence has been deposited in GISAID as hCoV-19/Russia/KLU-RII-MH107906S/2022 (EPI_ISL_15829108).

## Funding

This work has been supported by the RSF grant no 21-74-20160 to GVK.

## Acknowledgements

This work has been performed prior to April 2023. Sequencing has been supported by Russian Government subsidy for SARS-CoV-2 genomic sequencing (decree No. 267-r, 17.02.2022) to DAL.

## Notes

### Competing Interest Statement

The authors have declared no competing interest.

### Author Declarations

The research was approved by the Local Ethics Committee of Smorodintsev Research Institute of Influenza (protocol No. 203, 18 May 2023)

